# Dietary disparities of urban immigrant schoolchildren in New York City: Results from a mixed-methods pilot study

**DOI:** 10.1101/2020.05.01.20087486

**Authors:** Stella S. Yi, Neile K. Edens, Ashley Lederer, Janet Pan, Stella K. Chong, Jennifer A. Wong, Yan Li, Jeannette Beasley, Chau Trinh-Shevrin, Simona C. Kwon

**Affiliations:** NYU School of Medicine, Department of Population Health, New York, NY USA; Independent Consultant, Austin, Texas USA; Thoughtful Food Nutrition, New York, NY USA; The New York Academy of Medicine, New York, NY USA; Icahn School of Medicine at Mount Sinai, Department of Population Health Science and Policy, New York, NY USA; NYU School of Medicine, Department of Medicine, New York, NY USA

**Keywords:** Asian American, children, diet, sodium, obesity

## Abstract

**Background:** Obesity has been identified as an emerging health concern for Chinese American children; however, very little is known about diets in Asian American children.

**Objective:** To assess the feasibility of assessing diet of urban Chinese American children in an applied (school) setting and to gain insight on diet and drivers of dietary intakes from community nutrition experts.

**Design:** Data were from the Food Journal Project 2017, a school-based pilot study conducted by a multi-sector collaboration, and qualitative data from nutrition and community experts.

**Participants/Setting:** Children aged 8-12 (n=83) completed two dietary assessments using a food diary from January-June 2017. Children were then interviewed using the food diary as a guide; dietary data were entered into the ASA24 system by study staff. Chinese American children were compared to non-Chinese peers with respect to nutrient intake and the Healthy Eating Index 2010 (HEI-2010). Six semi-structured interviews and one panel discussion including two registered dietitians and community leaders with working knowledge of the Chinese American community were conducted from January-June 2018.

**Main Outcome Measures:** Nutrient intake and HEI-2010 scores.

**Statistical Analyses Performed:** Chi-squared and t-test comparisons, with statistical significance set at α=0.05.

**Results:** Adjusted for caloric intake, Chinese American children consumed 20% more sodium, 21% more protein but 27% less sugar compared to non-Chinese children. With regards to the HEI-2010, Chinese American children had less favorable whole grains and sodium scores; and more favorable seafood protein and empty calories scores compared to non-Chinese children. Qualitative data underscored the current burden of diet-related health disparities among Chinese American children and suggested Chinese American receptivity to family-based nutritional and cooking education interventions.

**Conclusions:** Sodium reduction and increasing whole grain intakes may be warranted in Chinese American children but should be verified with additional studies. Interventions to improve nutrition in this understudied population are critical.

**Research Snapshot:** *Research Question:* What are typical dietary intakes and current community and family-based challenges related to healthy eating in Chinese American children – an understudied disparity group?

*Key Findings:* Chinese American schoolchildren have high sodium and low whole grain intakes compared to non-Chinese peers. These specific dietary disparities work in concert with low physical activity levels and cultural norms in contributing to diet-related health disparities in this group. Family-based nutrition education and potential focus of dietetic counseling on sodium and whole grain intake areas are indicated to address these disparities.

## Introduction

Obesity has been identified as an emerging health concern for Chinese American children, particularly boys. According to recent data, one in four (21%) low income, urban Chinese American children are overweight or obese.^1^ Chinese American immigrant families face challenges unique to their community for maintaining healthy weight status owing to cultural factors and dietary norms; limited nutrition-related health literacy, and family/caregiver eating practices.^2^ Further, obesity prevalence in China is increasingly rapidly; in other words, obesity and related dietary behaviors may be less related to the acculturation process and more likely to already be present at the time of migration.^3^ Eating habits established during childhood persist through the life course, highlighting the need to promote healthy eating behaviors in young children.^4^

Despite this burden of obesity, very little is known about the diets of Asian American – let alone Chinese American children. A literature review on dietary practices in Asian American children identified 13 articles of varying methodological quality; only 4 included validated diet measurement.^5^ More recently, an analysis demonstrated that Asian American children in California consumed less fruits and vegetables compared to Non-Hispanic white peers.^6^ Analyses of dietary recall data from the National Health and Nutrition Examination Survey (NHANES) 2011-12 documented lower consumption of sugary drinks.^7^ Lower consumption of sugary drinks and unhealthy items (e.g., fried potatoes, fruit juice, fast food) in children were corroborated in another analysis, though patterns tended to differ by Asian subgroup (e.g., Chinese, Indian) – highlighting the necessity for disaggregated analyses by Asian subgroup.^8^ Beyond these examples, understanding of the diets in Asian American children or children of specific Asian American subgroups is extremely limited.

The objective of this paper was to assess feasibility of assessing dietary intakes of urban Chinese American children in an applied (school) setting and to and gain insight into drivers of dietary intakes from community nutrition experts. Nutrient intakes of Chinese American children were compared to intakes of their racially/ethnically diverse peer group from their schools. The hypothesis of this analysis was that unique nutrient intakes would be identified for Chinese American children.

## Materials and Methods

### Food Journal Project – Quantitative Data

Data for this analysis come from the Food Journal Project (FJP) 2017, a pilot and feasibility study conducted in collaboration between the NYU School of Medicine (NYU SOM), two New York City (NYC) elementary schools, and Common Threads – a national, nonprofit provider of cooking and nutrition education in low income schoolchildren. The primary objective of the FJP was to determine the feasibility of measuring dietary intake in schoolchildren using a validated method for dietary assessment: food diary-assisted 24 hour dietary recall.^9^ The FJP was conducted in 2 schools with cooking and nutrition education programming provided by Common Threads, which provides programs to schools where >80% of children receive free or reduced price lunch. According to publicly available data on schools at the NYC Department of Education website, 100% of the children attending our partner schools live in poverty.^10^ Prior to implementation of the FJP, NYU SOM study staff contacted and met with school wellness coordinators at each school – facilitated by Common Threads. Children participating Common Threads programming were invited to participate in the FJP; children were enrolled if they provided by parental consent and simple written assent. Given that children were participating in Common Threads throughout the school year and with variable timing with regards to data collection, we do not believe our results to be influenced by their program participation.

NYU SOM study staff developed key study materials and procedures, based on prior examples and published literature where possible; materials were tested in a small number of children and refined based on their feedback. The food diary was developed to be friendly and easy to follow by children in the target age group. Data collected included the time of day, where the food was consumed (e.g., home, school), with whom (e.g., mother, friend), where the food was prepared (e.g., home, restaurant), and the food and drink items and amounts. Interviewer paper materials including a script for performing multiple passes developed by a dietitian (AL),^11^ were accompanied by visual guides to aid children’s recall. Measuring cups and measuring spoons to probe amounts of food were offered if child could not remember amounts. To probe drink sizes, two different sets of pictures from the ASA24 were used. Children completed the food diary assessment at two time points, spaced approximately 6 months apart to capture the beginning and end of the school semester and both winter and late spring dietary patterns. Diaries were distributed over random weekend and weekdays and collected the next day; children were interviewed by trained NYU SOM study staff during school hours.

The ASA24 was used to facilitate data management. The ASA24 is a validated, publicly available, online 24-hour dietary recall tool developed by the National Cancer Institute. While the ASA24 was designed to be self-administered on a computer, a modified approach was utilized to maximize participation (i.e., maintaining brevity of interview) and validity of data, and for practical reasons (e.g., ASA24 requires internet which was not available at partnering school sites). A three step process was employed: 1) children kept a one day food diary; 2) children participated in a 24-hour dietary recall interview with study staff; and 3) data were entered into the ASA24 system by study staff. Study staff – which included 2 pre-dietary internship nutrition students - were trained by a dietitian (AL) on how to use the paper-based instrument in the schools, on interviewing, and on ASA24 data entry. Nutrient information was assessed using the Nutrition Data System for Research (NDSR). Additional data collected via survey (e.g., demographics, grade, overall diet quality, oral health) were entered and managed using REDCap electronic data capture tools hosted at the NYU-Health and Hospitals Corporation Clinical and Translational Science Institute.

Parent materials were translated into Spanish and Simplified Chinese as per request of the school coordinators. All study procedures and materials (English and translated) were approved by the NYU SOM Institutional Review Board (IRB) and the NYC Department of Education IRB.

### Provider Perspectives – Qualitative Data

To obtain provider perspectives, the research team (SSY, NKE, JAW) facilitated one-hour structured interviews with six community subject matter experts, including two registered dietitians, familiar with nutrition education in Chinese American children and families, and one two hour panel group discussion with all experts to gather clarity about the topic. Interviews were conducted in-person and over the phone as preferred by the participant. Participants provided verbal assent. Experts were purposively selected and invited via email from longstanding community-based organization partners focused on health disparities research and through existing relationships. Topic guide questions were developed collectively across study staff (SSY, NKE, JAW) and were based on prior literature and extant local knowledge of the community. All of the key informants participated in the facilitated expert panel discussion which drew from the themes of the interviews but focused on the role of nutrition education in mitigating disparities and action-oriented recommendations. The purpose of the panel discussion was to foster interactive discussion, share perspectives, and to identify resources and generate strategies to address nutrition-related disparities in Asian American children in NYC. Extensive written notes were collected from the semi-structured interviews; the panel discussion was transcribed from a video recording. The research team did not have prior experience in conducting qualitative research, but was overseen by an expert in qualitative research methods (SCK).

### Statistical Analyses

Data were pulled from REDCap and the ASA24 systems. We averaged the two dietary recalls per child which were conducted between the months of January through June 2017. Only those who performed two dietary recalls were included in the analysis. To identify Chinese American students from our sample, we utilized published lists of Chinese surnames.^12^ We stratified all analyses by race/ethnicity and by Chinese/non-Chinese. We utilize non-Chinese as the referent group (combining all others who are not Chinese including other Asian children) to maximize power for comparisons. However, because this may be viewed as a non-traditional approach for comparisons, we also ran data stratified by race/ethnicity. The results comparing Asian to non-Asian groups were similar to results comparing Chinese to non-Chinese groups, demonstrating the combined non-Chinese referent group was acceptable.

Demographic and other characteristics assessed at the first visit were summarized and compared between Chinese American and Non-Chinese children using chi-squared tests. The means of key nutrients and key nutrients per 1000 calories were assessed overall and by Chinese American ethnicity; differences in means were assessed using t-tests. The normalization to calories allows for a clearer understanding of nutrient intake differences since some nutrient intakes are highly correlated with caloric intake (e.g., sodium). The Healthy Eating Index (HEI) 2010 is a scale to measure diet quality relative to the 2010 Dietary Guidelines for Americans. This standardized tool has been determined to be a valid and reliable instrument for nutrition researchers. The HEI-2010 includes 12 components that sum to a possible maximum of 100 points. FJP participants’ HEI-2010 component and total scores and corresponding standard errors and 95% confidence intervals were calculated using macros created by the National Cancer Institute using the HEI Scoring Algorithm Method. All data were analyzed using STATA (v.12.1, College Station, TX). Statistical significance was set at p < 0.05.

The qualitative data was assessed using inductive content analysis.^13^ An initial codebook was created based on the topic guide and refined. The interview notes and panel discussion transcription were independently coded by two reviewers (SKC, JAW). Any discrepancies in coding were discussed with the study team until consensus was reached. Qualitative analysis findings were shared with participants to ensure that themes and concepts were correctly captured and coded.

## Results

### FJP Findings

Overall, the FJP was very well-received by children and they were enthusiastic to participate. Data collection and entry was facilitated by hiring interns with dietary data experience; prior familiarity with the ASA24-2016 may be critical as ‘best judgement calls’ needed to be made by interns for entry of some foods. Interestingly, the food diary may not be necessary as a tool for aiding recall, as others have concluded in 9-10 year old schoolchildren.^14,15^ Nearly half of our participating children did not have a food diary with them at the time of the recall, and upon comparison, key nutrients did not differ appreciably between those children that had the diary and those that did not. The food diary introduced complicated logistics for the school coordinator that could be avoided if it were not used. Additional implementation-related factors and practical lessons learned from the FJP experience are included in Table 1.

**Table 1.**
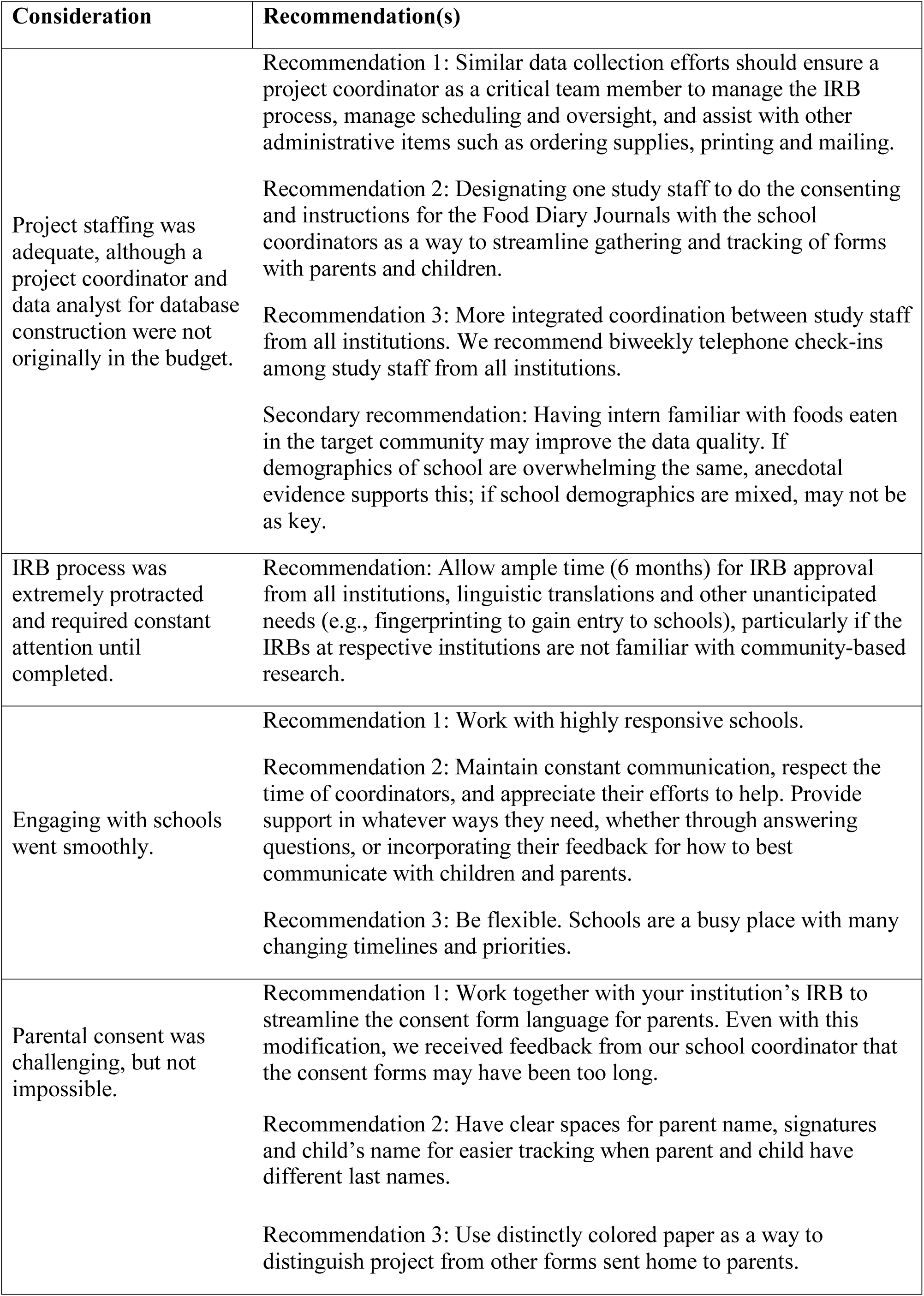
Implementation Considerations for Conducting Research in Urban Schools

A total of 94 children participated in the FJP. Three children did not perform a second dietary recall, and 8 children had race/ethnicity missing. The final analytic sample size for our analysis was n=83. Twenty-two children were identified as Chinese American, and 61 children were Non-Chinese: Hispanic (n=36), white (n=3), black (n=12) or Other Asian (n=10) (Table 2). Chinese American children did not differ from Non-Chinese children with respect to age, sex or grade.

**Table 2.** Demographics, Food Journal Project 2017, n=83

|  | <u>Overall</u> |  | <u>Chinese American</u> |  | <u>Non-Chinese</u> |  | <u>p-value*</u> |
| --- | --- | --- | --- | --- | --- | --- | --- |
|  | <u>n</u> | <u>%</u> | <u>n</u> | <u>%</u> | <u>n</u> | <u>%</u> |  |
| Age |  |  |  |  |  |  |  |
| 8 years | 13 | 15.7 | 2 | 9.1 | 11 | 18.0 | 0.57 |
| 9 years | 21 | 25.3 | 4 | 18.2 | 17 | 27.9 |  |
| 10 years | 39 | 47.0 | 13 | 59.1 | 26 | 42.6 |  |
| 11 years | 9 | 10.8 | 3 | 13.6 | 6 | 9.8 |  |
| 12 years | 1 | 1.2 | 0 | 0 | 1 | 1.6 |  |
| Sex |  |  |  |  |  |  |  |
| Male | 33 | 40.0 | 10 | 45.5 | 23 | 37.7 | 0.52 |
| Female | 50 | 60.0 | 12 | 54.6 | 38 | 62.3 |  |
| Grade |  |  |  |  |  |  |  |
| 3rd | 16 | 19.3 | 3 | 13.6 | 13 | 21.3 | 0.09 |
| 4th | 27 | 32.5 | 4 | 18.2 | 23 | 37.7 |  |
| 5th | 40 | 48.2 | 15 | 68.2 | 25 | 41.0 |  |
| Race/Ethnicity |  |  |  |  |  |  |  |
| Hispanic | 36 | 43.4 | 0 | 0 | 36 | 59.0 | <0.001 |
| White | 3 | 3.6 | 0 | 0 | 3 | 4.9 |  |
| Black | 12 | 14.5 | 0 | 0 | 12 | 19.7 |  |
| Asian | 32 | 38.6 | 22 | 100 | 10 | 16.4 |  |

\*Chi-squared test comparing Chinese American vs. Non-Chinese children

Mean values of key nutrients and key nutrients per 1000 calories are displayed in Table 2. Chinese American children consumed less sugars (29.6 less grams per day; p=0.01) compared to Non-Chinese children. When normalized per calories consumed, significant associations for higher sodium (306.8 mg/1000 kcal more, p<0.001) and protein (8.2 g/1000 kcal more, p=0.001) were observed; and persisted for lower sugar consumption (13.0 grams/1000 kcal, p=0.004).

The total HEI-2010 score for all children was 60.7, and did not differ significantly between Chinese American and Non-Chinese children (61.2 vs. 60.6, p=0.83; Table 3). The components of the HEI-2010, however, revealed interesting patterns. Chinese American children had less favorable whole grain (p=0.03) and sodium scores (p=0.004); and more favorable seafood and plant protein (p<0.001) and empty calories (p=0.02) scores compared to Non-Chinese children.

**Table 3.**
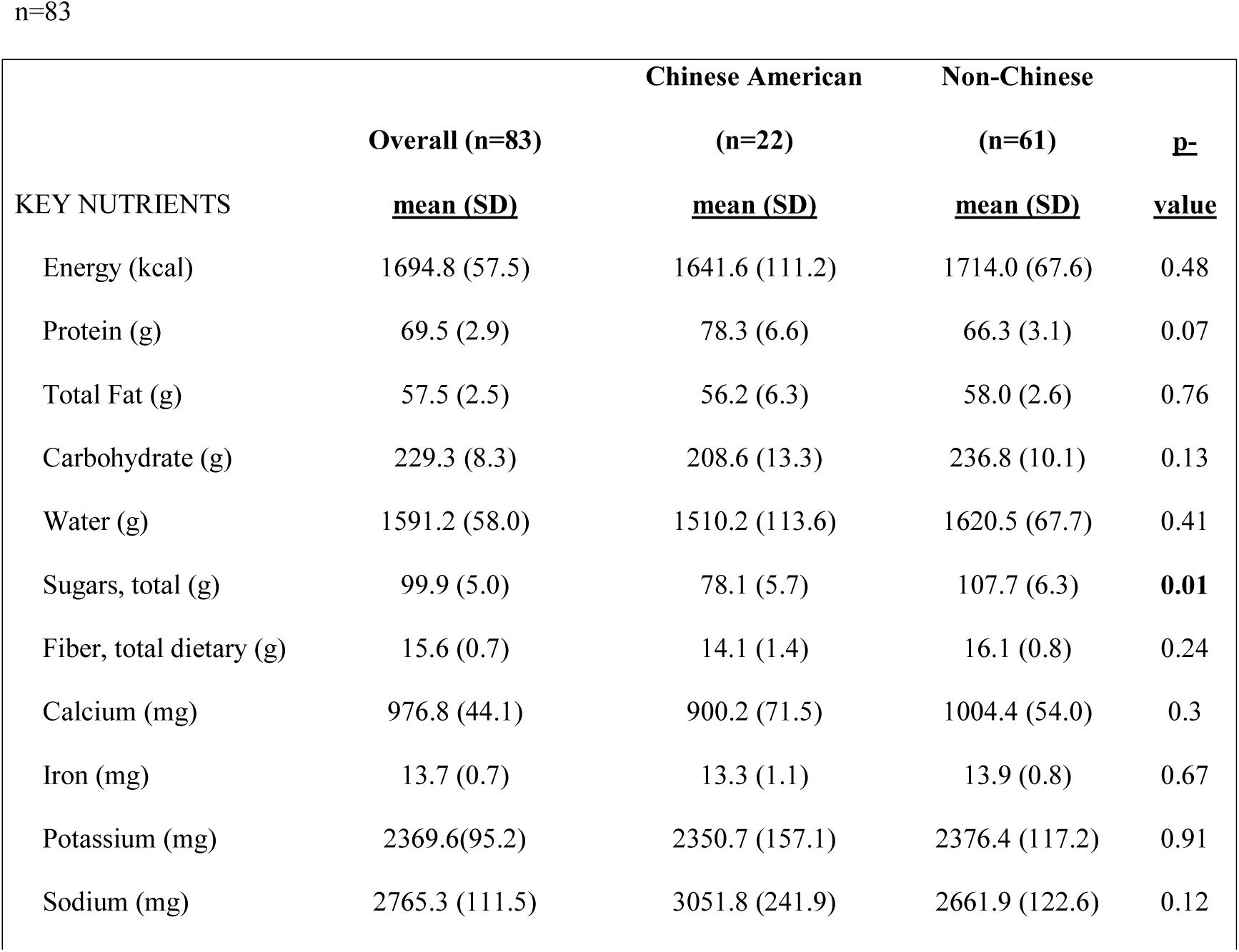
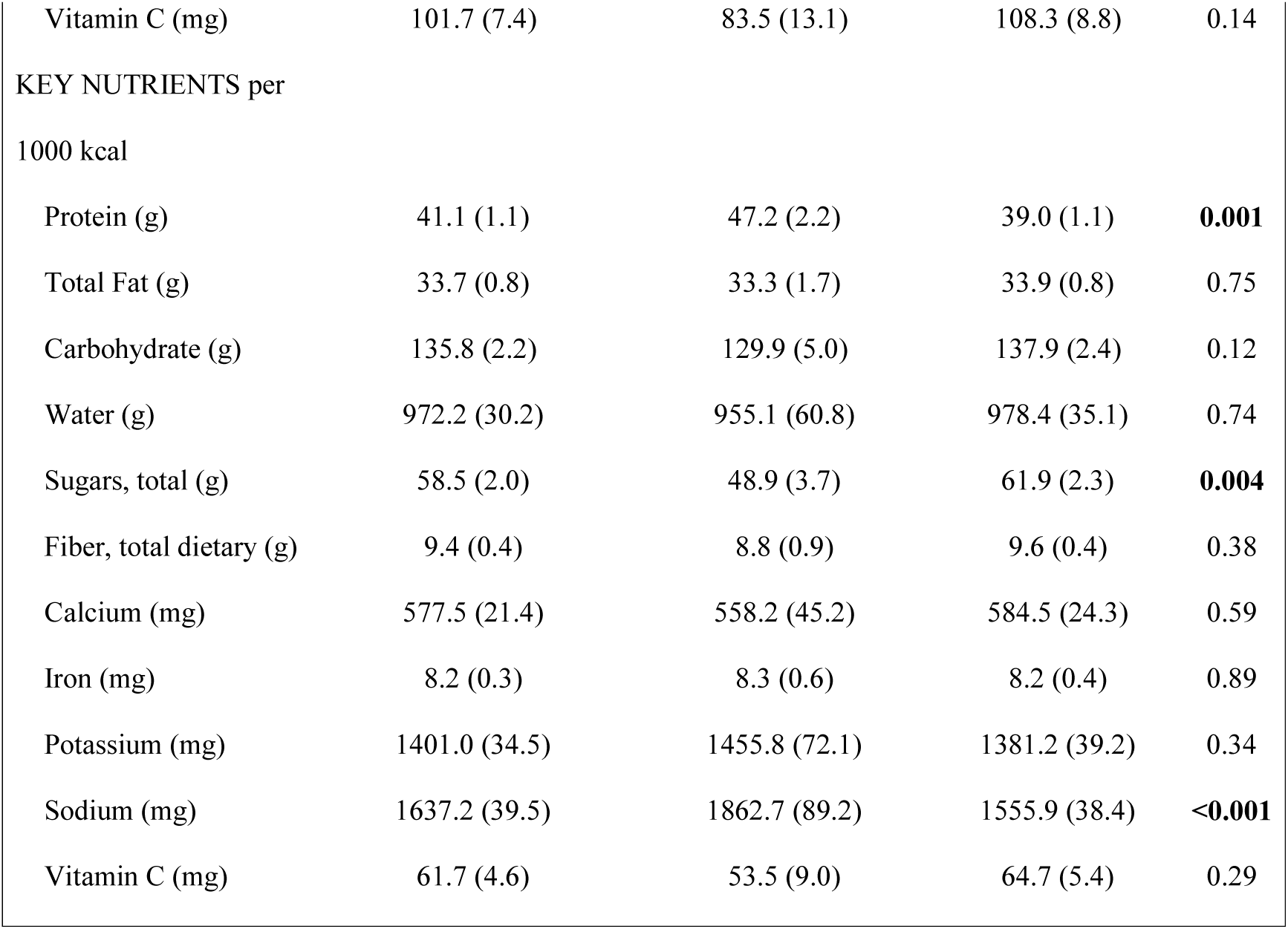
Mean Values of Key Nutrients and Key Nutrients Per 1000 Calories, Food Journal Project 2017

**Table 4.** Healthy Eating Index 2010, Food Journal Project 2017

|  | <u>Maximum score possible</u> | Overall (n=83)<br><u>mean</u> | Chinese American (n=22)<br><u>mean</u> | Non-Chinese (n=61)<br><u>mean</u> | <u>p-value</u> |
| --- | --- | --- | --- | --- | --- |
| **Adequacy (higher score, higher consumption)** |  |  |  |  |  |
| Hei-2010 Component 1 Total Vegetables | 5.0 | 2.4 (0.2) | 2.8 (0.4) | 2.3 (0.2) | 0.19 |
| Hei-2010 Component 2 Greens And Beans | 5.0 | 2.2 (0.2) | 2.3 (0.4) | 2.1 (0.3) | 0.70 |
| Hei-2010 Component 3 Total Fruit | 5.0 | 4.1 (0.2) | 3.6 (0.4) | 4.3 (0.2) | 0.06 |
| Hei-2010 Component 4 Whole Fruit | 5.0 | 4.1 (0.2) | 3.8 (0.4) | 4.3 (0.2) | 0.30 |
| Hei-2010 Component 5 Whole Grains | 10.0 | 3.6 (0.4) | 2.3 (0.5) | 4.1 (0.5) | **0.03** |
| Hei-2010 Component 6 Dairy | 10.0 | 7.4 (0.3) | 7.4 (0.6) | 7.4 (0.4) | 0.95 |
| Hei-2010 Component 7 Total Protein Foods | 5.0 | 4.2 (0.1) | 4.5 (0.2) | 4.1 (0.1) | 0.15 |
| Hei-2010 Component 8 Seafood And Plant Proteins | 5.0 | 2.4 (0.2) | 3.8 (0.4) | 1.9 (0.3) | **<0.001** |
| Hei-2010 Component 9 Fatty Acid Ratio | 10.0 | 4.6 (0.4) | 5.7 (0.7) | 4.2 (0.4) | 0.07 |
| **Moderation (higher score, lower consumption)** |  |  |  |  |  |
| Hei-2010 Component 10 Sodium | *10.0* | 4.4 (0.4) | 2.7 (0.7) | 5.0 (0.4) | **0.004** |
| Hei-2010 Component 11 Refined Grains | *10.0* | 5.2 (0.3) | 4.4 (0.7) | 5.5 (0.4) | 0.15 |
| Hei-2010 Component 12 Empty Calories | *20.0* | 16.0 (0.5) | 17.9 (0.8) | 15.4 (0.6) | **0.02** |
| Total HEI-2010 Score | *100.0* | 60.7 (1.3) | 61.2 (2.4) | 60.6 (1.5) | 0.83 |

Chinese American children reported more of the food they ate was sourced at school (27.5%) compared to Non-Chinese children (21.8%; p<0.01), although there was no difference in location that food was consumed (data not shown).

### Qualitative Data Findings

The semi-structured interview questions are displayed in Table 5. Several themes arose from the interviews and panel discussion that highlighted challenges to Chinese American children’s dietary consumption patterns seen in food journal data. These included:

#### Theme 1: High Levels of Sodium Consumption and Low Levels of Whole Grain Consumption

*“Perceptions of eating what you deem as cultural food, but they’re from the frozen food section. Recognizing that those foods are probably fried, high in* sodium.” (Local [NYC-based] public health dietitian, commenting on caregivers’ lack of knowledge related to sodium in processed foods; Panel)

**Table 5:**
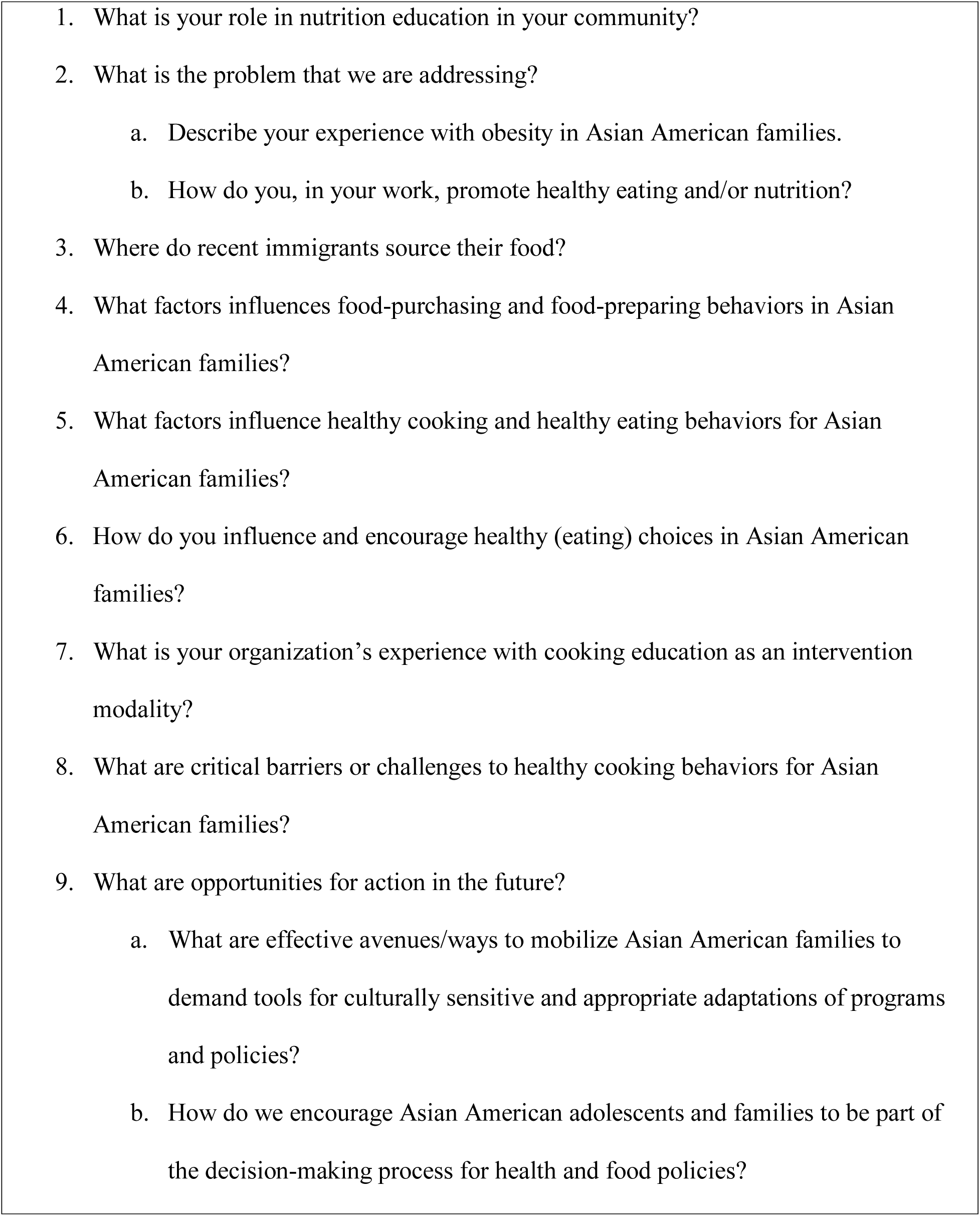
Interview Guide

**Table 6.** Nutrition Experts Recommended Calls to Action

| **Interviewed Expert** | **Organization** | **Call to Action** |
| --- | --- | --- |
| Administrator | NYC Department of Education (NYCDOE) | 1) Expand public outreach on health care and coverage for Asian immigrants;<br>2) Expand cultural competency: training for healthcare providers and revised healthcare services and materials;<br>3) Increase accessibility/affordability of physical activities for children;<br>4) Expand school-based interventions: utilize the school environment to educate children and families about nutrition via book drives and workshops for parents |
| Local (NYC-based)<br>Restaurateur/Chef-Owner |  | Celebrate food diversity and eat fewer processed foods, as much as possible. It is important to keep “plant-forward” food traditions alive not only to improve diet and health, but also to support sustainable practices for our planet. |
| Local [NYC-based] | NYC-based Private Nutritional | Offer cooking programs in schools and with the local |
| public health dietitian | Consultancy Firm | communities to encourage dietary behavioral shifts and to empower children to learn more about foods and have new experiences that they can bring to their parents and use for the rest of their lives. |
| Director | NYC Chinese American community-based organization | 1) Engage families and teachers in dialogue about food, culture, and health.<br>2) Offer nutrition and cooking classes at schools to involve teachers and families. |
| Dietitian | Federally Qualified Health Center (FQHC) serving primarily Chinese American immigrant community in NYC | 1) Provide nutrition education to children and to caregivers, particularly to grandparents. 2) Emphasize importance of family and community engagement in physical activity. |
| Pediatrician | Federally Qualified Health Center (FQHC) serving primarily Chinese American immigrant community in NYC | Increase breastfeeding education in Chinese American community to emphasize the health benefits to children's development and growth, and as a means of preventing obesity. |
| Academic Researcher | NYC Research University | 1) Increase participation of Asian Americans in public health |
|  |  | research to reduce Asian American health disparities.<br>2) Raise awareness and share knowledge about Asian American health disparities among researchers and local communities to collaboratively prevent Asian American health disparities. |

Interview and panel respondents reported observing high sodium and refined grains (white rice, noodles) consumption in Chinese American children’s diets – particularly in after school/study center settings. Respondents also associated this phenomenon with Chinese American children’s parents and grandparents being unaware of high sodium present in traditional Chinese dishes and in processed foods frequently incorporated into dishes, such as canned meats or preserved vegetables. Additionally, interviewees reported that Chinese American communities are generally unfamiliar with brown rice preparation, that brown rice is not readily available in local markets – and when available, is more expensive. Respondents’ feedback about the difficulty in persuading Chinese Americans to adopt whole grain in their diets is in line with our local and others work concerning the stigma of brown rice consumption.^16,17^

#### Theme 2: Grandparents’ Role on Children’s Dietary Intake

*“Grandparents only want to give [their grandchildren] what they didn’t have.”*

(Director from Chinese American community-based organization, commenting on grandparents’ reasons for yielding to grandchildren’s food requests, Panel)

Respondents observed that grandparents are increasingly the main caregivers of grandchildren, responsible for preparing meals before or after school, oftentimes for the entire family. Grandparents thus have significant influence on grandchildren’s dietary intake, supporting existing literature describing an association between grandparental care and childhood overweight and obesity.^18–20^ Interview respondents noted that grandparent caregivers often recall their personal struggles with poverty and experiences with limited food resources. These recollections may motivate their overindulgence and overfeeding in response to their grandchildren’s food requests.

#### Theme 3: Significant Cultural Identity of Food

*“Grandparents feel like a meal is not a meal if they don’t eat rice. The word in Chinese for rice is the same as a meal – so I have to clarify, do you mean they don’t eat rice or do you mean they don’t eat dinner.”* (Pediatrician from FQHC, Panel)

*“We are getting an influx of younger parents into our school system, so we are trying to educate our parents that the things we may have grown up with, are not being done right… Those were the things we grew up learning and passed onto our students and our children. These are things we have to debunk and demystify.”* (NYC DOE Administrator, commenting on the dietary and behavioral challenge posed by the cultural significance of food in Chinese American households, Panel)

Cultural identity is extremely important in Chinese American families, influencing the type of food prepared and served to children at home as a way to uphold cultural values. Chinese Americans’ dietary familiarity with traditional staple foods such as rice and noodles make it challenging to alter or to eliminate these starch-heavy foods with whole grain options from diets. Interviewees also described grandparents preparing or consuming “special occasion foods” and encouraging their children and grandchildren to eat them as a form of sharing cultural identity, despite low nutritional value of these foods. The phenomenon of increased consumption of special occasion or festival foods among immigrant groups has been previously documented in the literature as one explanation for increased cardio-metabolic risk in these populations.^21^

#### Theme 4: Increased Consumption of Unhealthy Junk Foods

*“As they go to elementary and middle school, the kids adapt to the American diets and lifestyles, even though in our culture we eat a lot of fruits and vegetables. However, they adapt to the American diet, eating things like pizza, chips and soda.”* (Dietitian from FQHC, Panel)

Interviewees observed that traditional Chinese culture is less influential in school-age children’s individual food preferences and choices. Many Chinese American children and adolescents assert preference for more Americanized foods, which are often processed and less healthy.

#### Theme 5: Oral Health Disparities

*“It’s always a challenge to convince a caregiver to see the dentist… It’s because in China they don’t have a preventative dental system…”* (Pediatrician from FQHC, Panel)

Experts expressed concern over the prevalence of oral health disparities they observe among Chinese American children, attributed to a lack of preventative dental care education in China and high consumption of culturally normative sugar-sweetened beverages, like bubble teas and fruit juices in the US. Disparities in oral health in this community have been identified in ours and others’ prior research.^22,23^

#### Theme 6: Lack of Physical Activity

*“I asked them, “Do you get any kind of physical activity? “ And they say they do not, they just go to daycare to make sure their homework is done. Physical activity is very limited in our population, because they see that academics is more important. “* (Dietitian from FQHC, Panel)

Respondents observe a lack of physical activity among Chinese American children and adolescents. Parents place greater emphasis on academic extracurricular programs than on physical after-school activities, like team sports. Respondents noted that many caregivers lack the time and supplemental income to send their children to gyms or exercise programs like swim lessons. We and others have previously documented Asian Americans to have low levels of physical activity,^24–27^ while other prior research corroborates the emphasis of academic vs. exercise norms in Chinese American children.^2^

#### Theme 7: Nutritional/Cooking Education as an Effective Intervention

*“It’s a lot about educating the parents. We know [they] want to spoil them, but [they] also have to understand that they have to make better choices for them.”* (Director from Chinese American community-based organization, Panel)

*“The parents and students were not really English proficient, but cooking became a common language for them. We had an event where all the parents came out and they shared their cooking skills throughout the year. The students were so proud to be able to come out and serve the parents what they had made.”* (NYC DOE Administrator, Panel)

Interview respondents believe that nutritional education programs targeting parent and grandparent caregivers, and also teachers, may be effective interventions to help improving understanding of healthy food choices, behaviors, and practices and reduce overweight and obesity in Chinese American children. Combined nutritional/cooking education can engage parents and students to cook and to learn about new foods together, breaking any language barriers and bonding parents and children. Hands-on nutritional and cooking classes empower students and parents to try new foods and be more aware of the types of foods and nutritional values of the foods they consume. This idea has also been reflected in additional data we have collected in the Chinese American adult community, where the majority respondents supported nutrition education (70%) as a potential mechanism to improve the diets of community members.^28^

## Discussion

We found that it was feasible to assess the diets of urban, immigrant Chinese American schoolchildren in underserved areas aged 8-12 years using food diary-assisted 24-hour dietary recalls and compared them to their peers. To our knowledge, this is the first comprehensive characterization of diets in Chinese American children in the United States using a validated method. We found that Chinese American children in our sample consumed more sodium dense (mg per 1000 calories) diets and less sugar compared to Non-Chinese children. Using HEI scores, we also identified that Chinese American children had less favorable whole grain intakes, and more favorable seafood-and-plant-protein and empty calories intake compared to Non-Chinese children. The sodium and sugar findings are consistent with prior literature using less comprehensive measures; while the differences in protein sources and lower whole grain intakes have not been previously characterized. In Asian American (not Chinese specifically) adults, sodium density has also been shown to be higher compared to other racial/ethnic groups.^29,30^ The higher seafood consumption is reflective of prior U.S. data on Asian Americans 6 years and older,^31^ an aspect of diet which is both positive and negative. Higher seafood consumption offers favorable fatty acid benefits, but is also associated with high exposure to neurotoxic heavy metals and higher sodium intake in Asian Americans.^32^ Both high sodium intake and heavy metals increase the risk of subsequent health conditions, and are therefore of concern.

Chinese American immigrants are an underserved population. National and local data demonstrate that Chinese Americans have similar poverty rates as other racial/ethnic minority groups (national poverty rate: Chinese American – non-citizen immigrant, 26%; white, 11%; black, 26%; Hispanic, 24%; ^33^ NYC poverty rate, Chinese Americans, 21%; white, 14%; black, 22%; Hispanic, 25%).^34^ Yet broad racial stereotypes both societally and in the research community that this community suffers from few health disparities^35^ have contributed to limited knowledge of dietary behaviors^36^ and a lack of nutrition-related interventions in this group. This lack of data is troubling given that Asian American adults and children are increasingly at risk for development of cardio-metabolic disorders – including diabetes,^37,38^ prediabetes^39^ and non-alcoholic fatty liver disease^40–42^ – despite lower estimates of overweight and obesity compared to other racial/ethnic groups.^43^

Our findings complement the prior literature demonstrating that Chinese American families face unique dietary challenges upon immigration to the U.S.^2^ First, because of a lack of familiarity with the American diet and ingredients, Chinese immigrant parents may be less able to provide nutritious meals and health information to their children compared to U.S. born parents.^2^ After attending school, many children of Chinese immigrants tend to develop preferences for American food, driven in part by peer influence, and these preferences may lead to familial (intergenerational) conflict when the children requests unfamiliar American food that is disliked by the parents.^2^ When dietary accommodations are made to appease the children (a common approach in Chinese culture),^44^ the American foods incorporated are often convenience foods that are unhealthy (e.g., desserts, salty snacks, high-fat meat).^45^ Second, many Chinese American immigrants reside in low income, urban enclaves, where convenience versions of traditional foods high in fat and sodium are readily available.^46^ Asian foods, even unhealthy versions, are perceived to be healthier than American foods and are often chosen.^21^ Third, dietary acculturation practices tend to involve the incorporation of unhealthy vs. healthy American foods (e.g., convenience foods, salty snacks).^45^ Fourth, unhealthy dietary practices are introduced by grandparents.^2,20^ Chinese grandparents introduce belief systems of appropriate body size (plumpness) in children, and may have potentially experienced early life deprivation leading to overindulgence with food for their grandchildren.^2,20^ Lastly, owing to the model minority stereotype and low community and provider awareness about the nature of obesity risk for Asian American populations, Chinese American parents and caregivers may perceive their risk to be low and be less likely to seek help or clinical interventions^2^ or to refer patients. Compounding this issue are structural challenges, such as limited resources and services for Chinese American families facing issues around obesity, including culturally appropriate materials, information or programs. This directly points to the need for increased awareness and education among registered dietitians to offer appropriate assessments and services to these children and their families.

A strength of the current manuscript is the demonstration of the feasibility of conducting a somewhat complex method of data collection within a school setting and among an ethnically diverse student body. While data collection was recommended to be in English by the school coordinators among the students we worked with, this fact may not be generalizable to other immigrant children. Further strengths include the presentation of analyses focused on a specific subgroup of Asian Americans using a validated and comprehensive method for assessing diet. Asian Americans are often not included in national or regional surveys, and were only recently added to NHANES in the 2011-12 survey wave, therefore very little is known about diet in Asian American adults or children. We also use two rather than one dietary recall, strengthening both the validity and reliability of our results. The limitations of this analysis include that data were collected from the children themselves, which may be influenced by social desirability, intrusion, or forgetfulness. Food products such as types of bread (e.g., whole wheat vs. partial wheat) and dairy (e.g., 1% vs. 2% milk) that were not visually distinctive were particularly difficult. We note that this may be the case particularly for empty calories, as our study participants seemed to anchor their recall around meals (breakfast, lunch and dinner). We specifically prompted for recalls of treats or snacks, but we would caution the interpretation of the empty calories score for this reason. Our nutrient/food specific results may be generalizable to children of similar demographic profile (i.e., largely immigrant, low income). For example, our interns reported that “every child consumed white rice” – which may not be applicable to the general U.S. child population where the predominant carbohydrates are bread products. We were unable to take into account changes in the menus at the school cafeteria, though this effect would be somewhat mitigated by having two time points per child. While we feel that the food diary/ASA24-2016 was feasible, we’d be remiss to not mention the limitations of the ASA24-2016 and any dietary recall data in general – though not a limitation of the FJP per se. In particular, the ASA24-2016 method may have limitations in qualifying ethnic foods.

The qualitative data findings generated possible reasons for the observed differences in dietary intake between CA and other children, as well as suggestions for possible intervention approaches to address overweight and obesity among Chinese American children and adolescents. Experts’ calls to action are summarized in Table 5.

The results of our analysis provide a preliminary understanding of diets among Chinese American children, and highlight the need for larger cross-sectional and longitudinal descriptive studies as well as potential avenues for future intervention work. Specific interventions focusing on sodium reduction, increasing whole grain consumption, potentially replacing animal with vegetable protein sources, and monitoring of heavy metals in Chinese American children may be warranted. A deeper understanding of other dietary behaviors, particularly around sugar intake and specific ethnic products may be key. Interventions to educate Chinese American immigrant parents and grandparents about the significance of overweight and obesity in their community and healthy dietary habits may also be critical in improving diets amongst these children. Focus group findings suggest that a multifaceted approach in addressing overweight and obesity among Chinese American children is necessary. Both school-based and environmental based interventions, specifically nutritional and cooking education are strongly advocated by experts.

## Data Availability

Data may be requested by establishing a Data Use Agreement with the lead author/lead author organization.

## Acknowledgements

Thanks are due to the staff and leadership at Common Threads for facilitating researcher outreach to schools, and for moderating/co-hosting the panel discussion. We also thank the school administrators, teachers, and students who participated in the project, and the expert community panelists who participated in the qualitative portion of this study.

